# Covid-19: analysis of a modified SEIR model, a comparison of different intervention strategies and projections for India

**DOI:** 10.1101/2020.06.04.20122580

**Authors:** Arghya Das, Abhishek Dhar, Srashti Goyal, Anupam Kundu

**Affiliations:** International Center for Theoretical Sciences, Tata Institute of Fundamental Research, Bangalore-560089, India

## Abstract

To understand the spread of Covid-19, we analyse an extended Susceptible-Exposed-Infected-Recovered (SEIR) model that accounts for asymptomatic carriers, and explore the effect of different intervention strategies such as social distancing (SD) and testing-quarantining (TQ). The two intervention strategies (SD and TQ) try to reduce the disease reproductive number, *R*_0_, to a target value 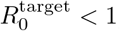, but in distinct ways, which we implement in our model equations. We find that for the same 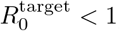, TQ is more efficient in controlling the pandemic than SD. However, for TQ to be effective, it has to be based on contact tracing and our study quantifies the required ratio of tests-per-day to the number of new cases-per-day. Our analysis shows that the largest eigenvalue of the linearised dynamics provides a simple understanding of the disease progression, both pre- and post-intervention, and explains observed data for many countries. We propose an accurate way of specifying initial conditions for the numerics (from insufficient data) using the fact that the early time exponential growth is well-described by the dominant eigenvector of the linearized equations. Weak intervention strategies (that reduce *R*_0_ but not sufficiently) reduce the peak values of infections and the asymptotic affected population and we provide analytic expressions for these in terms of the disease parameters. We apply them in the Indian context to obtain heuristic projections for the course of the pandemic, noting that the predictions strongly depend on the assumed fraction of asymptomatic carriers.

---

The Covid-19 pandemic, that started in Wuhan (China) around December 2019, has now affected almost every country in the world. The total number of confirmed case on May 30 was close to 5.9 million with close to 360, 000 deaths. One of the serious concerns presently is that there is as yet no clear picture or consensus on the future evolution of the pandemic. It is also not clear as to what the ideal intervention strategy that a government should implement, while taking into account also economic and social factors. The role of mathematical models has been to provide guidance for policy makers [1–13].

One of the standard epidemiological model is the SEIR model which has four compartments of susceptible (*S*), exposed (*E*), Infected (*I*) and Recovered (*R*) individuals with *S* + *E* + *I* + *R* = *N* being the total population of a region (the model can be applied at the level of a country or a state or a city and is expected to work better for well-mixed populations). The SEIR model is parameterized by the three parameters *β*, σ and *γ* that specify the rates of transitions from *S* → *E, E* → *I* and *I* →*R* respectively. In terms of the data that is typically measured and reported, *R* corresponds to the total number of cases till the present date, while *γI* would be the number of new cases per day. The number of deaths across different countries is some fraction (≈ 1−10%) of *R* [18] while the number of hospital beds required at any time would be ≈ (new cases per day × typical days to recovery). An important parameter characterizing the disease growth is the reproductive number *R*_0_ — when this has a value greater than 1, the disease grows exponentially. Typical values reported in the literature for Covid-19 are in the range *R*_0_ = 2 − 7 [14]. For the SEIR model one has *R*_0_ = *β/γ*.

The two main intervention schemes for controlling the pandemic are social distancing (SD) and testing-quarantining (TQ). Lockdowns (LD) impose social distancing and effectively reduce contacts between the susceptible and infected populations, while testing-quarantining means that there is an extra channel to remove people from the infectious population. These two intervention schemes have to be incorporated in the model in distinctive ways [4, 5] — SD effectively changes the infectivity parameter *β* while TQ changes the parameter *γ*. Intervention schemes attempt to reduce this to a value less than 1. In the context of the SEIR model with *R*_0_ = *β/γ*, it is clear that we can reduce *R*_0_ by either decreasing *β* or by increasing *γ*. In this work we point out that for the same reduction in *R*_0_ value, the effect on disease progression can be quite different for the two intervention strategies.

Here we analyze intervention strategies in an extended version of the SEIR model which incorporates the fact that asymptomatic or mildly symptomatic individuals [4–6, 15] are believed to play a significant role in the transmission of Covid-19. Our extended model considers eight compartments of Susceptible (*S*), Exposed (*E*), asymptomatic Infected (*I*_*a*_), presymptomatic Infected (*I*_*p*_), and a further four compartments (*U*_*a*_, *D*_*a*_, *U*_*p*_, *D*_*p*_), two corresponding to each of the two infectious compartments. These last four classes comprise of individuals who have either recovered (at home or in a hospital) or are still under treatment or have died — they do not contribute to spreading the infection (see Sec. (III A) for details). We do not include separate compartments for the number of hospitalized and dead since these extra details would not affect our main conclusions. For this extended SEIR model we discuss the performance of two different intervention strategies (namely SD and TQ) in the disease dynamics and control. We study both the cases, of strong interventions (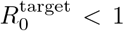 aimed at disease suppression,) and that of weak interventions (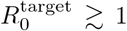, aimed at disease mitigation).

## Main conclusions

**Apart from the reproductive number**, *R*_0_, **an important parameter is the largest eigenvalue of the linear dynamics, which we denote as** *µ*. **For** 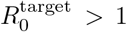, **we have** *µ* > 0 **and this gives us the exponential growth rate (doubling time** ≈ 0.7*/µ***). On the other hand, for** *R*^target^ < 1, **the corresponding** *µ* **is less than** 0 **and this tells us that infections will decrease exponentially. For the case where only mitigation is achieved** 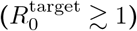, **we present analytic expressions for peak infection numbers, time to reach peak values, and asymptotic values of total affected populations. These provide useful guidance on disease progression and we apply it in the Indian context. We also show that, for the same reduction of** 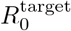 **to a value less than** 1, **the corresponding** *µ* **magnitude can be very different for different intervention schemes. A larger magnitude of** *µ*, **corresponding to a faster suppression of the pandemic, is obtained from TQ than that from SD. We give conditions for TQ to be successful: (a) it has to be based on contact-tracing and (b) it is necessary that testing numbers are scaled up according to the number of new detected cases. We show that the above picture gives us a comprehensive understanding of data from several countries which have either achieved disease suppression or mitigation**.

### A note on the Indian situation

*The number of daily new cases in India continues to rise and it is clear that only mitigation has been achieved, unlike in Europe and the US which have succeeded in suppression* (*R*_0_ < 1). *The disease doubling time is around* 14 *days* (*as on May 20*). *For two different choices of the fraction of asymptomatics* (*and typical values of disease parameters*) *our estimates suggest a value of* 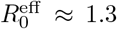 *and that the disease would peak between July to September. The predicted number of hospitalizations and deaths per day* (*assuming* 1% *deaths for symptomatic cases*) *have a large uncertainty but could be quite large* [*see Tables* (*I-III*)], *and there is an urgent need of preparing for this. However, the lockdown in India is now being eased. Given the huge economic and social costs of implementing hard SD, it is clear that a combination of weaker SD but intense TQ might be the only practical way of controlling the pandemic in India. A sustained and targeted testing and quarantining strategy* (*assuming community spreading is still limited*), *combined with some level of social-distancing has to be implemented at the earliest and to the fullest extent. Community transmission is unlikely to have taken place in all states and cities in India. Increasing the testing-to-detected ratio to a value around* ≳ 100 *from the value of* ≈ 25 (*as on May 15*) *could result in lowering of R*_0_ *and the µ value or at least in not letting them increase further*.

The rest of the paper is structured as follows. In Sec. I we state our main results. In Sec. I C we make comparisons of the predictions of the SEIR model with real data on confirmed number of cases and make some heuristic predictions in the Indian context. We summarize our results in Sec. II. All the technical details including the precise definition of the extended SEIR model with and without interventions and our analytic results are given in Sec. III.

## I. RESULTS

The extended SEIR model studied here is schematically described in Fig. 1. It has eight variables (*S, E, I*_*a*_, *I*_*p*_, *U*_*a*_, *D*_*a*_, *U*_*p*_, *D*_*p*_) and ten parameters (*β*_*a*_, *β*_*p*_, *σ, γ*_*a*_, *γ*_*p*_, *α, ν*_*a*_, *ν*_*p*_, *r, u*), of which *α* represents the fraction of asymptomatic carriers while *u, r, ν*_*a*_, *ν*_*p*_ are related to intervention strategies. The model details are given in Sec. III A. For the present, we note that at any given time the total infectious population size is *I* = *I*_*a*_ + *I*_*p*_, the cumulative affected population (recovered, in hospital or dead) is *R* = *U*_*a*_ + *D*_*a*_ + *U*_*p*_ + *D*_*p*_, the reported total confirmed cases is *C* = *D*_*a*_ + *D*_*p*_ + *U*_*p*_, and the reported new daily cases is *D* = *dC/dt* = *rν*_*a*_*I*_*a*_ + (*γ*_*p*_ + *rν*_*p*_)*I*_*p*_.

**FIG. 1.**
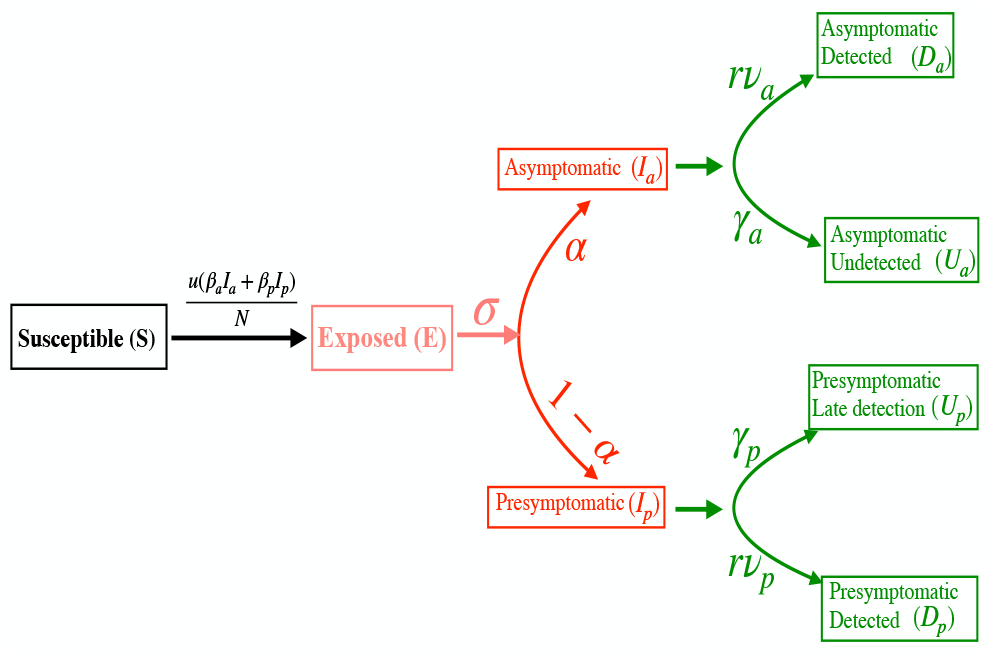
A schematic description of the extended SEIR dynamics studied in this work. The parameters *β*_*a*_, *β*_*p*_, *σ, γ*_*a*_, *γ*_*p*_, *α* are intrinsic to the disease, *u* quantifies the degree of social distancing while *ν*_*a*_, *ν*_*p*_, *r* are related to intervention arising from testing-quarantining.

The parameter *u* quantifies the degree of social distancing while *r* is related to the rate at which testing-quarantining is done. These are in general time-dependent, *u* changing from the free (without interventions) value *u* = 1 to a target value *u*_*l*_ < 1, while *r* is a rate that changes from 0 to a value *r*_*l*_ > 0. The time-scale for the change depends on how efficiently the control measures are implemented.

### A. Comparison of different intervention strategies

A useful quantity to characterize the system with interventions is the targeted reproduction number [see Eqs. (43,44)]

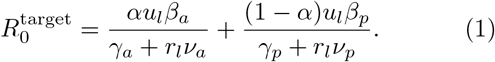

We classify intervention strategies by the targeted 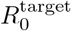 value. A **strong intervention** is one where 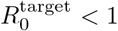 and will achieve suppression of the disease while a **weak intervention** is one with 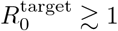 and will only mitigate the effects of the disease.

Other than *R*_0_, an important quantity to characterize the disease growth is the largest eigenvalue *µ* of the linearized dynamics (see Sec. III B). In the early phase of the pandemic, all populations other than *S* grow exponentially with time as ∼ *e*^*µt*^. As we will see, for the case of strong intervention, *µ* becomes negative and gives the exponential decay rate of the disease.

In our numerical study we choose, for the purpose of illustration, the following parameter set: *α* = 0.67 and the rates *β*_*a*_ = 0.333, *β*_*p*_ = 0.5, σ= 1*/*3, *γ*_*a*_ = 1*/*8, *γ*_*p*_ = 1*/*12 all in units of day^−1^. For the specified choice of parameter values (free case with *u* = 1.0, *r* = 0.0) we get *µ* = 0.158 which is close to the value observed for the early time data for confirmed cases in India. The corresponding free value of *R*_0_ is 3.7665. Note that *µ* is not uniquely fixed by *R*_0_ (and vice versa) and different choices of parameters can give the same observed *µ* but different values of *R*_0_

Choosing these typical parameter values for Covid-19, we now compare the efficacy of strong and weak interventions implemented in four different ways: (1) 6WLD-NTQ: Six weeks lockdown (strong value of SD parameter) and no testing-quarantining, (2) ELD-NTQ: Extended lockdown and no testing-quarantining,(3)NSD-ETQ: No social distancing and extended testing-quarantining, (4) ESD-ETQ: Extended social distancing and extended testing-quarantining. The case with no social distancing and no testing-quarantining is indicated as NSD-NTQ.

We work with a population *N* = 10^7^ and initial conditions *E*(0) = 100, *I*_*a*_(0) = *I*_*p*_(0) = *U*_*a*_(0) = *D*_*a*_(0) = *U*_*p*_(0) = *D*_*p*_(0) = 0 and *S*(0) = *N* −*E*− *I*_*a*_− *I*_*p*_− *U*_*a*_− *D*_*a*_− *U*_*p*_ −*D*_*p*_. In all cases, we will assume that intervention strategies are switched on when the confirmed number of cases reaches 50 and after that the full intervention values are attained over a time scale of 5 days.

#### 1. Strong intervention *(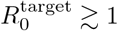)*

In this case, the exponential growth stops around the time when 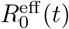 crosses the value 1. After this time, the infection numbers will start decaying exponentially. Since the infection numbers are still small compared to the total population, one can work with the linearized theory and the magnitude of the largest eigenvalue *µ* (now negative) determines the exponential decay rate. For illustrating this case, we take:

##### Parameter set I 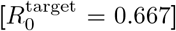

We choose three SD and TQ strengths as (i) SD: *u*_*l*_ = 0.177, *r*_*l*_ = 0, TQ: *u*_*l*_ = 1, *r*_*l*_ = 1.2 and (iii) SD-TQ: *u*_*l*_ = 0.461, *r*_*l*_ = 0.4. This choice corresponds to changing the free value of *R*_0_ = 3.766 to a target value = 0.667, 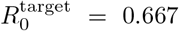 for all the three different strategies. The largest eigenvalue *µ* changes from the free value *µ* = −0.158 to the values (i) *µ*− = 0.027, (ii) *µ*− = 0.077 *µ* =− 0.0546 respectively. The results of the numerical solution of the extended SEIR equations are presented Figs. (2a) and (2b).

### Main observations

1. A six week (or eight week) lockdown is insufficient to end the pandemic and will lead to a second wave. If the interventions are carried on indefinitely, the pandemic is suppressed and only affects a very small fraction of the population (less than 0.1%). We can understand all features of the dynamics from the linear theory. In Figs. 2(a,b), intervention is switched on after ≈ 2 weeks and the peak in infections appears roughly after a period of 5 days. Thereafter however, the decay in the number of infections occurs slowly, the decay rate being given by the largest eigenvalue *µ* (now negative and smaller in magnitude than *µ* in the growth phase).
2. *We find that for the same target* 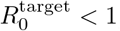, *different intervention schemes* (*ELD-NTQ, NSD-ETQ, or ESD-ETQ*) *can give very different values of the decay rate µ and, in general we find that TQ is more effective than SD*. We see that ELD-NTQ ends the pandemic in about 10 months while NSD-ETQ would take around 3.5 months. This can be understood from the fact that the corresponding *µ* values (post-intervention) are given by *µ* − = 0.027 and *µ*−= 0.077 respectively, i.e, they differ by a factor of about 3. With a mixed strategy where one allows almost three times more social contacts (*u*_*l*_ = 0.431) than for LD case and that requires three times less testing (*r*_*l*_ = 0.4) than for TQ case, we see that the disease is controlled in about 5 months. Hence this appears to be the most practical and effective strategy.
3. The expected time for the pandemic to die would be roughly given by

**FIG. 2.**
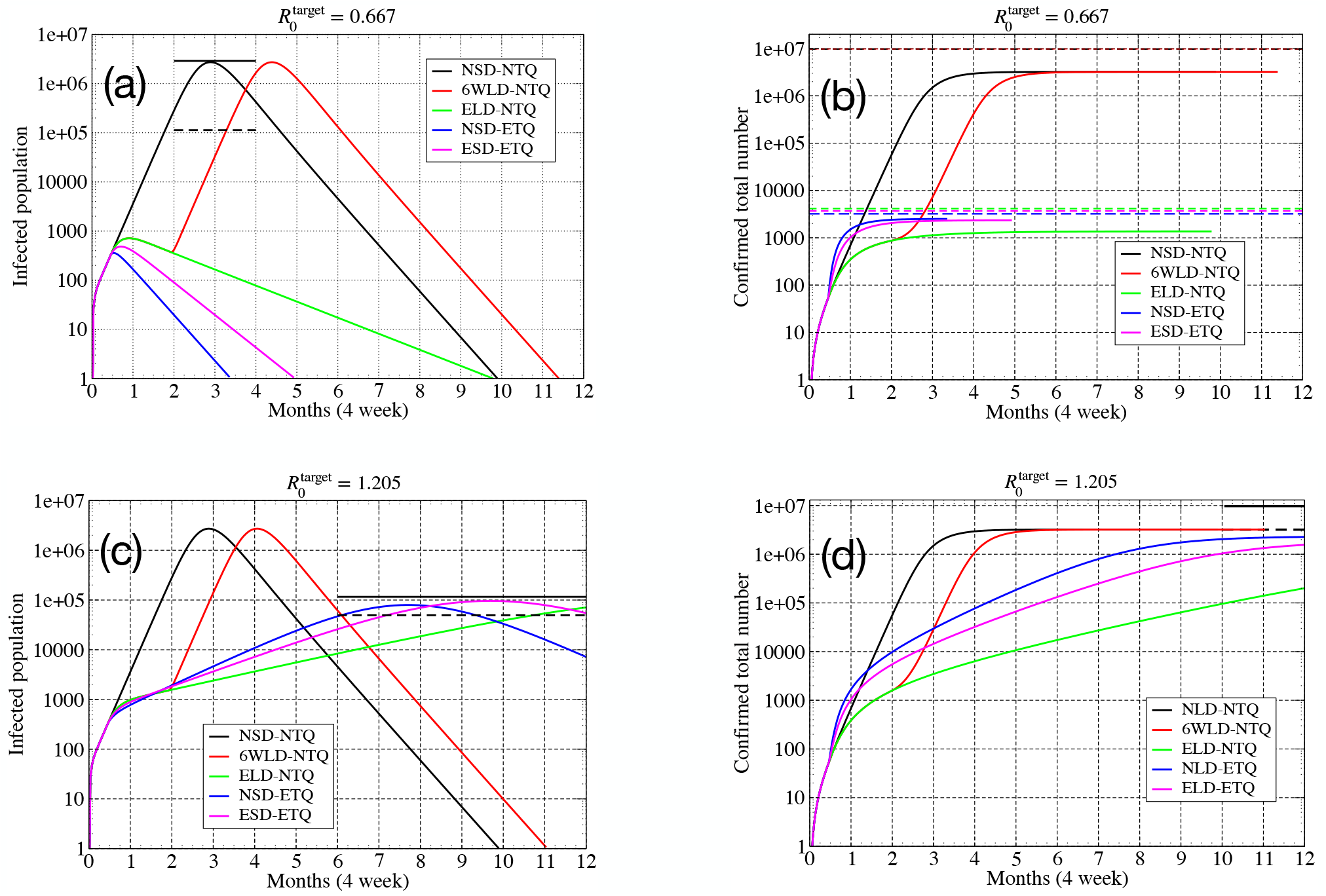
**Parameter set I** 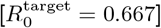: (a) Total number of infected cases *I* = *I*_*a*_ + *I*_*p*_ for different intervention strategies. The solid and dashed black lines indicate the peak infected cases (*I*^(*m*)^) as given by (3) and the corresponding value of 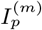. (b) Total number of confirmed cases *C* = *U*_*p*_ + *D*_*a*_ + *D*_*p*_. The dashed lines indicate the total affected population *R* = *C* + *U*_*a*_ at the end of one year, for the different strategies. In the absence of interventions this is close to 96% and is given by (5). The total population was taken as *N* = 10^7^. **Parameter set II** 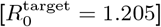: (c) Total number of infected cases *I* = *I*_*a*_ + *I*_*p*_ for different intervention strategies. (d) Total number of confirmed cases *C* = *U*_*p*_ + *D*_*a*_ + *D*_*p*_. The dashed lines indicate the total affected population *R* = *C* + *U*_*a*_ at the end of one year, for the different strategies. Total population was taken as *N* = 10^7^.

**FIG. 3.**
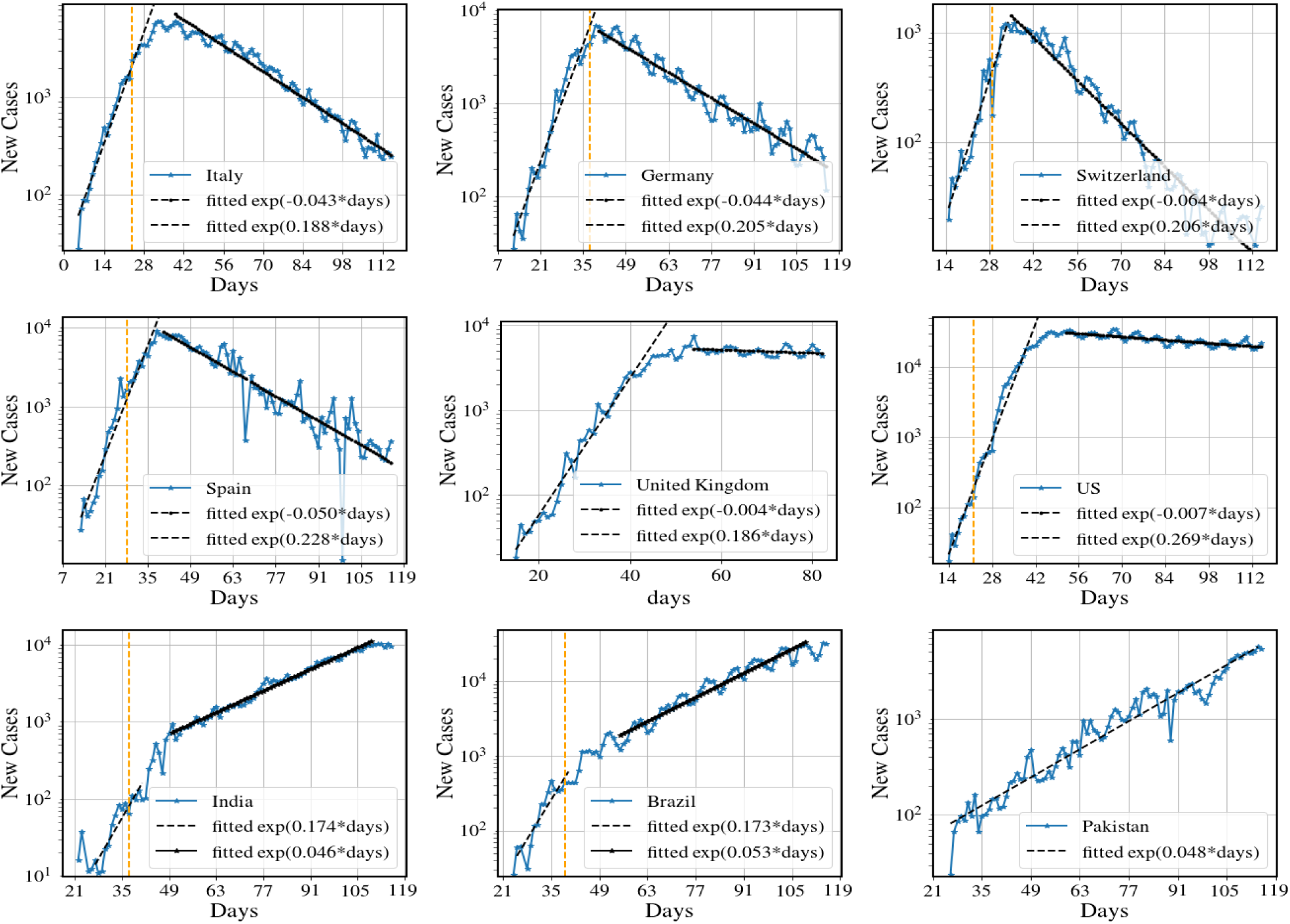
Number of new cases per day for nine different countries. We note that the first six data sets exhibits the same broad features that we see for the model predictions in Fig. 2(a,b). In particular we see the fast exponential growth and slow exponential decrease in new cases (following strong interventions). The two countries UK and US show a very slow decay rate, indicating that disease suppression has barely been achieved. The data for India, Brazil and Pakistan show the behavior corresponding to model predictions in Fig. 2(c,d) and have only been able to achieve mitigation so far 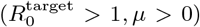. Data from [19] and the end date is June 10.

and so it is important that intervention schemes are implemented early and as strongly as possible.

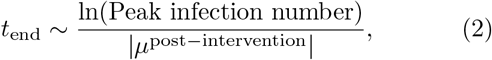

#### 2. Weak intervention 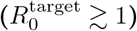

In this case, a finite fraction of the population is eventually affected, but the intervention succeeds in reducing this from its original free value and in delaying considerably the date at which the infections peak. We take the following parameter set for this study:

##### Parameter set II 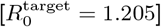

we choose three SD and TQ strengths as (i) SD: *u*_*l*_ = 0.32, *r*_*l*_ = 0, (ii) TQ: *u*_*l*_ = 1, *r*_*l*_ = 0.536 and (iii) SD-TQ: *u*_*l*_ = 0.634, *r*_*l*_ = 0.24. This choice corresponds to changing the free value of *R*_0_ = 3.766 to a fixed target value 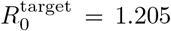 for all the three different strategies. The largest eigenvalue *µ* remains positive and changes from the free value *µ* = 0.158 to the values (i) *µ* = 0.0152, (ii) *µ* = 0.032 (iii) *µ* = 0.0248 respectively. The results of the numerical solution of the extended SEIR equations are presented Figs. (2c) and (2d).

### Main observations

1. *We find that in this case the peak infections, peak infection time and the final affected population can be obtaied from analytic expressions in terms of µ, R*_0_ *and a few other basic disease parameters*. One can use these formulas either using the pre-intervention or post-intervention values of *R*_0_ and *µ*. Assuming that we start with a small seed infected or exposed population and with almost the entire population susceptible, i.e *S*(0) ≈ *N*, the peak value of infections, *I*^(*m*)^, (which is proportional to the number of hospitalizations required) and the number of days, *t*^(*m*)^, to reach this peak value are given by the simple general relations

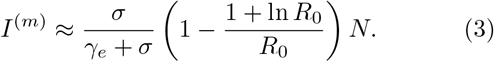

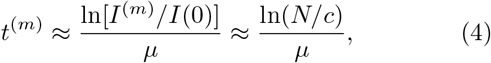

where *γ*_*e*_ is an effective recovery rate (see Eq. 39 in Sec. III C] and *c* is a constant that depends on initial infected population and other disease parameters. The fraction of population, 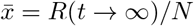, that is eventually affected is given by the solution of the equation

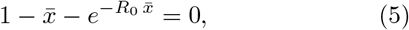

this result being valid for very general SEIR-type models with multiple compartments. We also provide relations for estimating the number of asymptomatic infected and recovered individuals. These analytic results are very useful to obtain quick heuristic estimates for the typical numbers for peak infections and for the time when the peak occurs. In Sec. I C we use these to make predictions for India, which is an example where only mitigation 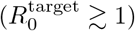 has been achieved.
2. We find that the peak infection numbers are smallest for the case with ELD-NTQ and they occur at a later stage. These results can also be understood mathematically from the expressions in (3) and (4) using the post-intervention values of *γ* and *µ* (from the linear theory).
3. We note that while weak interventions can slow down and reduce the impact of the pandemic, they do not lead to development of herd immunity of the population (assuming that all the recovered people develop immunity). It is well known that herd immunity is attained when a fraction 1−1*/R*_0_ of the population has developed immunity. Thus herd immunity in the above example would require that 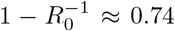 i.e 74% of the population be affected, while (5) with 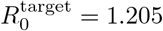 predicts that only about 31% of the population is affected.

### B. Results from the linearized theory

As already observed, the linear theory is very useful in understanding the growth and also the decay time scales of the pandemic following strong interventions. Another observation that we make is that, independent of initial conditions, the vector describing all the system variables quickly points along the direction of the eigenvector corresponding to the largest eigenvalue. Hence if we know any one variable (or a linear combination of all the variables) at sufficiently large times in the growing phase, then the full vector is completely specified. *This leads to an accurate way of specifying initial conditions for the numerics* (*from insufficient data*) and will help in reducing the number of fitting parameters in modeling studies, thereby increasing their accuracy in predicting. This fact also implies that different initial conditions (such as different seed infections) will only cause a temporal shift of the observed evolution. Plotting the data for number of confirmed cases (normalized by the population), starting with the same initial value, should therefore lead to a collapse of the data for different countries. We test this idea and find that indeed an approximate collapse of data is obtained for a number of countries (see next section).

### C. Comparisons with observed data for Covid-19

In this section we discuss a comparison of our results with real data on evolution of the Covid pandemic in different countries. We do not attempt a detailed comparison of the model predictions with the data since there are too many poorly known parameters and possibly quite inaccurate knowledge of the initial conditions of the variables themselves. Rather we make some overall qualitative observations relating data to the predictions from SEIR-type models and find that in many cases, several broad qualitative features are remarkably well captured by the model. For the case of India we discuss predictions, based on our analytic formulas, on the disease evolution for a range of choice of parameter values.

#### 1. Observation of strong and weak intervention in Covid-19 data

In Figs. (3) we give some examples of data for number of new cases for nine countries where we see that some of the qualitative features seen in the model results in Fig. 2(a,b). In particular we see the fast exponential growth phase and then a much slower decay phase for the first six countries which have succeeded in controlling the disease with various levels of success. On the other hand we see that India, Brazil and Pakistan continue to show a positive *µ* and it is clear that intervention schemes need to be strengthened.

One issue is that different countries start with different initial conditions (for example the seed exposed population could be very different between countries). As discussed in Sec. III B, as long as the number of confirmed cases is much smaller than the population size, a description in terms of the linearized dynamics is accurate. This would predict an initial exponential growth and then as intervention schemes begin to operate, the reproductive number and the corresponding growth exponent would decrease till eventually one is able to achieve 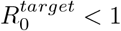 and correspondingly *µ* < 0. In Fig. 5 we show data for the reported number of new cases in 12 different countries and approximately see these features. Most countries have succeeded in disease suppression 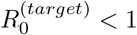, but show a slow exponential decay of the disease. A few Asian countries (India, Pakistan, Indonesia) have not yet entered the decaying phase which means that intervention has been weak and only disease mitigation has been achieved. This means that with the same level of intervention strategy, a finite fraction of the population will eventually be affected in these countries. We discuss later in some more detail the Indian situation.

**FIG. 4.**
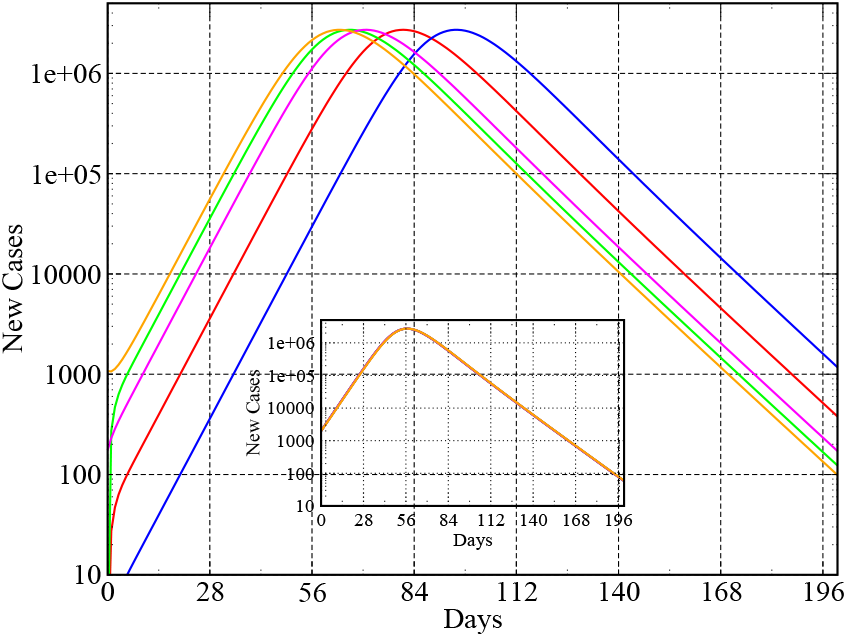
**Role of initial conditions**: Plot showing *I*(*t*), for a fixed population of size *N* = 10^7^, with 5 very different initial conditions : (1)*E*(0) = 100, *I*_*a*_(0) = 0, *I*_*p*_(0) = 0, (2)*E*(0) = 10, *I*_*a*_(0) = 0, *I*_*p*_(0) = 0, (3)*E*(0) = 1000, *I*_*a*_(0) = 0, *I*_*p*_(0) = 0, (4) *E*(0) = 233, *I*_*a*_(0) = 100, *I*_*p*_(0) = 75, (5) *E*(0) = 233, *I*_*a*_(0) = 1000, *I*_*p*_(0) = 75. (Inset) A collapse of all the curves obtained by translating all the trajectories so that they start with the same value of *I*.

**FIG. 5.**
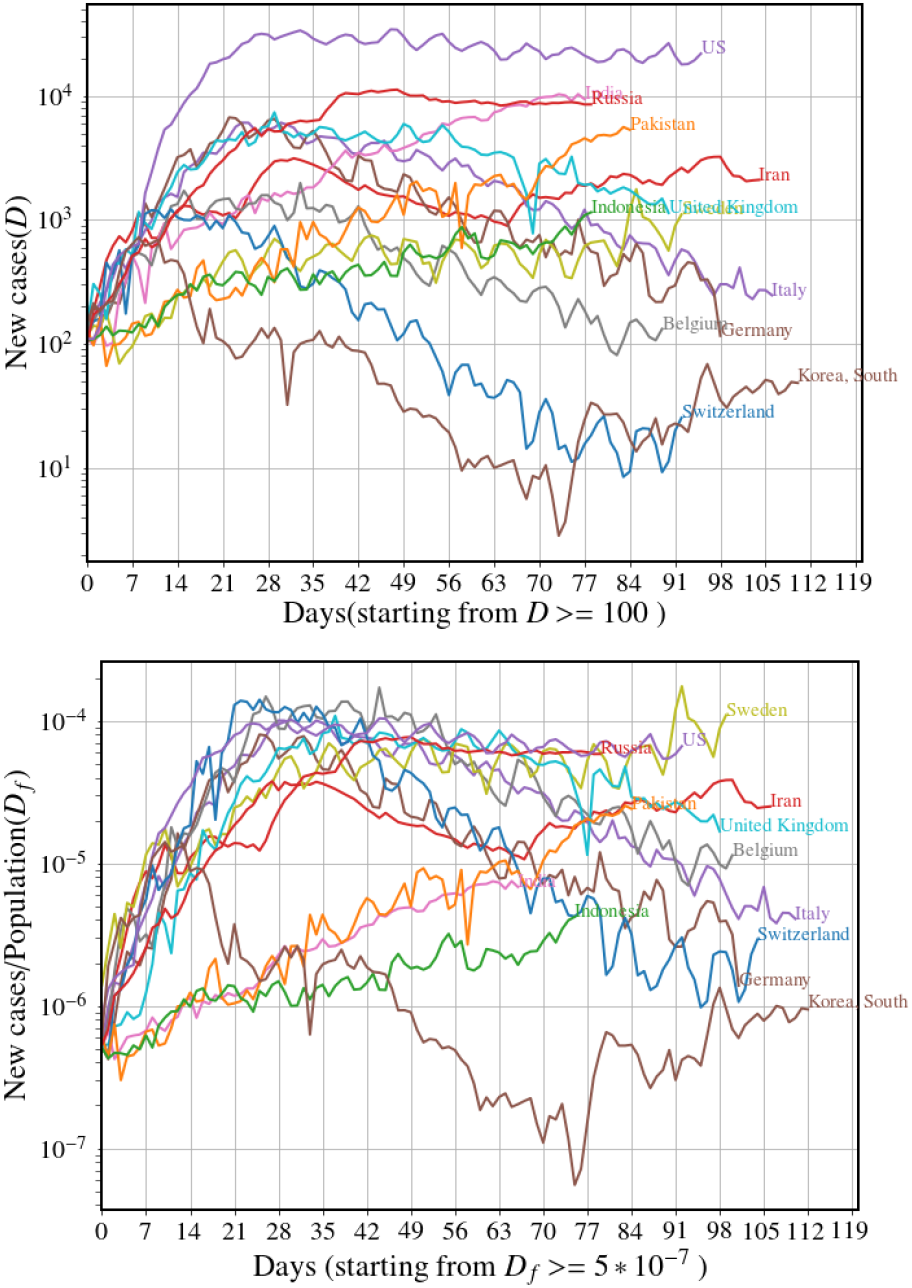
(left) Number of new cases per day for different countries. (right) Number of new cases normalized by the total population, with the time axis shifted so that every country starts with the same normalized value. Data from [19].

#### 2. Comparing data across different countries

The linearized SEIR dynamics also predicts that (see Sec. III B), if one uses similar parameters and intervention parameters, then all countries should follow the same trajectory provided they start with the same value for the normalized fraction of confirmed new cases (*D*_0_*/N*). We illustrate this idea, for the extended SEIR dynamics, in Fig. 4 where we show a plot of *I*(*t*) = *I*_*a*_(*t*) + *I*_*p*_(*t*) for 5 different initial conditions. The inset shows a collapse of all the trajectories after an appropriate time translation of the different trajectories. Can we see a similar collapse of the real data for different countries (after normalizing by the respective populations and with appropriate time translation of the data) ? In the right panel of Fig. 5 we plot the data with this normalization and initial condition and see a rough collapse for several countries. We notice in particular that three of the Asian countries (India, Pakistan, Indonesia) follow a distinctly different trajectory — this could indicate either that the disease parameters are different or that the intervention strategies have been different, or the reporting of cases is inaccurate.

### D. How much testing is required?

How is *r* related to testing rates ? It is easy to see that with random testing of the population, intervention can be helpful only if a finite fraction of the population is tested, which is practically very difficult to implement. Thus testing has to be based on contact tracing of the new detected cases. Suppose that the number of tests per day is *T* while the number of daily new cases is Δ*D* and *A* the typical number of contacts made by a person over the period when the person is infectious and before detection. Following arguments described in Sec. (III E) and Eqs. (45-46), we find that TQ intervention can be successful only if one achieves

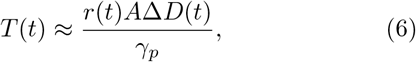

where *r*(*t*), our control rate function changes from the value 0 to a value *r*_*l*_, which should be at least of the same order as *γ*_*p*_, the recovery rate of symptomatic carriers. This means that we need *T* (*t*) ≈ *A*Δ*D*(*t*), that is *the number of tests/per day has to be proportional to number of new detections/per day and in fact the ratio T/*Δ*D has to be larger than the average number of contacts, A, that each infectious person makes*. The number *A* is expected to depend on the population density and also how well SD is being implemented. Hence, while the value of *T* (*t*)*/*Δ*D*(*t*) ≈ 25 (around May 15) for India appears to be large, it may not be sufficient given that the population densities are much larger than in many other countries and implementation of SD may be less effective. If we assume 20 contacts a day and the number of days before isolation of the individual to be 5 we get the rough estimate of *A*≈ 100 and then the ratio *T/*Δ*D* thus has to be at least ≈ 100. This is the minimum value of testing-to-detected ratio that has to be targeted at localities with high infection rates. The details of our arguments, described later (see Sec. (III E)), are largely *independent of the specifics of the particular SEIR model that we study*.

### E. Predictions for India from extended SEIR model

In the following we make some heuristic predictions, based on the analytic results in Eqs. (3-5) and the present observed data, for daily new cases in India (*N* ≈ 1.3 ×10^9^), in the state of Delhi (*N* ≈ 1.9 ×10^7^) and in the city of Mumbai (*N*≈ 1.3× 10^7^). The analysis here is based on the assumption of a best case scenario where the value of 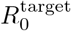 (achieved after a nationwide lockdown of 6 weeks) will be maintained.

Here we assume that intervention has effectively been through SD, with *r* << *γ* being neglected. We consider the following choice of parameter values which appears to be reasonable for getting a conservative estimate: 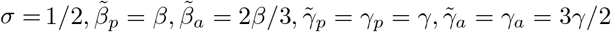, i.e, we assume that asymptomatics are 2*/*3rd less infectious and recover 3*/*2 times faster. This gives us [using (39)] *γ*_*e*_ = *γ/*(1 − *α/*3) and the effective reproductive number as 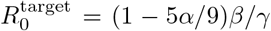. From this last relation we can write 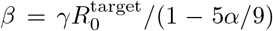. Plugging this into the equation for the eigenvalues, (19), and replacing *λ* by the observed mean exponential growth rate *µ* ≈ 0.05 (the value observed for India since around April 10, see Fig. 3), we see that we basically get an equation for 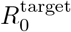 in terms of *α, σ, γ* and *µ*. For specific choices of *α*, σ and *γ*, the observed values of *µ* before and after intervention will then give us the corresponding values of *R*_0_.

**TABLE I.** Predictions for India with different choices of parameter values.

| $\sigma$ | $\gamma$ | $\alpha$ | $R_0$ (free) | $R_0^{\text{target}}$ | Peak daily cases (PDC) | Time of peak | Total affected | Total deaths |
| --- | --- | --- | --- | --- | --- | --- | --- | --- |
| 0.5 | 0.2 | 0.67 | 2.28 | 1.33 | 2,456,630 | 2nd week September | 45 % | 1,936,000 |
| 0.5 | 0.2 | 0.9 | 2.16 | 1.3 | 686,770 | 3rd week August | 42 % | 550,700 |
| 0.4 | 0.143 | 0.67 | 2.82 | 1.45 | 2,956,600 | 3rd week September | 55 % | 2,363,700 |
| 0.4 | 0.143 | 0.9 | 2.65 | 1.41 | 832,890. | 4th week August | 52 % | 676,140 |

**TABLE II.** Predictions for Delhi with different choices of the asymptomatic fraction *α*.

| $\sigma$ | $\gamma$ | $\alpha$ | $R_0$ (free) | $R_0^{\text{target}}$ | Peak daily cases (PDC) | Time of peak | Total affected | Total deaths |
| --- | --- | --- | --- | --- | --- | --- | --- | --- |
| 0.5 | 0.2 | 0.67 | 2.28 | 1.33 | 35,904 | 1st week August | 45 % | 28,296 |
| 0.5 | 0.2 | 0.9 | 2.16 | 1.3 | 10,037 | 2nd week July | 42.3 % | 8,048 |
| 0.4 | 0.143 | 0.67 | 2.82 | 1.45 | 43,212 | 2nd week August | 55 % | 34,546 |
| 0.4 | 0.143 | 0.9 | 2.65 | 1.41 | 12,173 | 2nd week July | 52.3 % | 9,882 |

**TABLE III.** Predictions for Mumbai with different choices of the asymptomatic fraction *α*.

| $\sigma$ | $\gamma$ | $\alpha$ | $R_0$ (free) | $R_0^{\text{target}}$ | Peak daily cases (PDC) | Time of peak | Total affected | Total deaths |
| --- | --- | --- | --- | --- | --- | --- | --- | --- |
| 0.5 | 0.2 | 0.67 | 2.28 | 1.33 | 24,566 | 3rd week July | 45 % | 19,360 |
| 0.5 | 0.2 | 0.9 | 2.16 | 1.3 | 6,867 | 4th week June | 42.3 % | 5,507 |
| 0.4 | 0.143 | 0.67 | 2.82 | 1.45 | 29,566 | 3rd week July | 55 % | 23,637 |
| 0.4 | 0.143 | 0.9 | 2.65 | 1.41 | 8,328 | 4th week June | 52 % | 6,761 |

For our analysis we need to know the total infections *I*(0) on some day (we take this as April 11) and we estimate it in the following way. Suppose that the daily observed cases on this day was *D*_*p*_(0) (assuming that only the symptomatics are detected). Then we have *I*_p_ (0) = *D*_p_ (0)*/γ*. From (40) we have 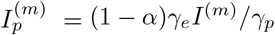, and so the time to the peak can be estimated as 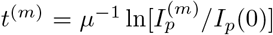. We use Eq. (3) to compute the peak number of infections *I*^(*m*)^ and the peak daily cases (PDC) is then obtained as 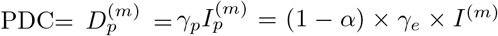. The total affected population fraction 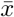, can be computed from Eq. (5), using only the knowledge of 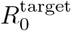. If we assume the number of deaths is 1% of all symptomatic cases this gives us an estimate for the total number of deaths as 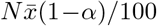.

The observed daily new cases in India, Delhi and Mumbai on April 10 were around *D*_*p*_(0) ≈ 900, *D*_*p*_(0) ≈ 115 and *D*_*p*_(0) ≈ 195 respectively [20]. For a range of choice of the parameters with σ = 0.5, 0.4, *γ* = 0.2, 0.143 and of *α* = 0.67,0.9, we compute the corresponding values obtained for *R*_0_ and 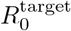. These and the estimates for 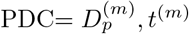 and 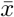 are given in Tables I, II and III for India, Delhi and Mumbai. Note that while the peak numbers and total affected population and deaths simply scale with population size, the time to peak depends on the daily detected numbers on April 10, and this leads to the observed differences in the time to the peak for the three cases. We also note here that changing the initial conditions by about 10% causes a change of few days in the peak time while the other quantities remain unchanged. The full numerical solution [see Fig. (9) in Sec. (III)] also shows that the complete suppression of the disease takes more than 6 months after the peak.

We point out that the mixed-population assumption the SEIR model is expected to be more accurate for a smaller population and so the estimates for Delhi and Mumbai would be more reliable than the one for India. For a big and highly in-homogeneous country like India, smaller regions (states or cities) would have different values of *µ* and *R*_0_ and also different initial conditions, hence the global values would not capture the local dynamics correctly. It is likely that the numbers in Table I are an over-estimate of the true future trajectory. For the state of Delhi and the city of Mumbai these would be more accurate, *however we see that the uncertainty in the true value of α leads to a huge uncertainty in the predictions*.

## II. DISCUSSION

In summary a modified version of the SEIR model, incorporating asymptomatic individuals, was used for analyzing the effectiveness of different intervention protocols in controlling the growth of the Covid-19 pandemic. Non-clinical interventions can be either through social distancing or through testing-quarantining. Our results indicate that a combination of both, implemented over an extended period may be the most effective and practical strategy. We point out that short-term lock-downs cannot stop a recurrence of the pandemic if interventions are completely relaxed and developing herd immunity is not a practical solution either since this would affect a very large fraction of the population.

We have provided numerical examples to illustrate the basic ideas and in addition, have stated a number of analytical results which can be useful in making empirical estimates of various important quantities that provide information on the disease progression. Looking at real data for new Covid-19 cases in several countries, we find that the extended SEIR model captures some important qualitative features and hence could provide guidance in policy-making. We use our analytic formulas to make predictions for disease peak numbers and expected time to peak for India, the state of Delhi and the city of Mumbai, but point out that these predictions could be incorrect for India (due to big inhomogeneity in disease progression across the country) and perhaps more reliable for the cases of Delhi and Mumbai. Our formulas are easy to use and give quick heuristic estimates on disease progression, which would be reliable when applied to local populations (in towns, cities and perhaps smaller countries). While the lack of precise knowledge of the disease parameters (e.g the fraction of asymptomatic carriers) leads to rather large uncertainties in the predictions, they could perhaps be used to obtain reasonable bounds.

## Data Availability

All data are from public data bases which have been cited.

## III. METHODS

### A. Definition of the extended SEIR model

We consider a population of size *N* that is divided into eight compartments:

1. *S* = Susceptible individuals.
2. *E* = Exposed but not yet contagious individuals.
3. *I*_*a*_ = Asymptomatic, either develop no symptoms or mild symptoms.
4. *I*_*p*_ = Presymptomatic, those who would eventually develop strong symptoms.
5. *U*_*a*_ = Undetected asymptomatic individuals who have recovered.
6. *D*_*a*_ = Asymptomatic individuals who are detected because of directed testing-quarantining, may have mild symptoms, and would have been placed under home isolation (few in India).
7. *U*_*p*_ = Presymptomatic individuals who are detected at a late stage after they develop serious symptoms and report to hospitals.
8. *D*_*p*_ = Presymptomatic individuals who are detected because of directed testing-quarantining.

We have the constraint that *N* = *S* +*E* +*I*_*a*_ +*I*_*p*_ +*U*_*a*_ + *D*_*a*_ + *U*_*p*_ + *D*_*p*_. A standard dynamics for the population classes is given by the following set of equations:

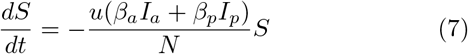

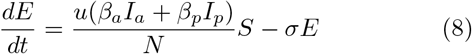

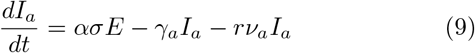

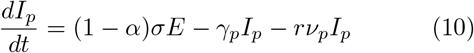

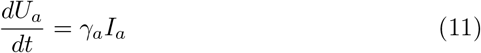

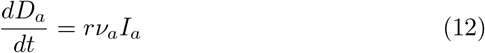

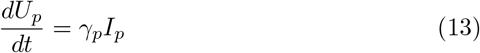

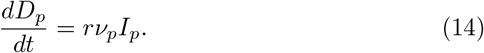

The parameters in the above equation correspond to

- *α*: fraction of asymptomatic carriers.
- *β*_*a*_: infectivity of asymptomatic carriers.
- *β*_*p*_: infectivity of presymptomatic carriers.
- *σ*: transition rate from exposed to infectious.
- *γ*_*a*_: transition rate of asymptomatic carriers to recovery or hospitalization.
- *γ*_*p*_: transition rate of presymptomatics to recovery or hospitalization.
- *ν*_*a*_, *ν*_*p*_: detection probabilities of asymptomatic carriers and symptomatic carriers. Here we choose *ν*_*a*_ = 1*/*3, *ν*_*p*_ = 1*/*2,
- *u*: intervention factor due to social distancing (time dependence specified below).
- *r*: intervention factor due to testing-quarantining (time dependence specified below). This is a rate and depends on testing-quarantining rates.

With our definitions, the total number of confirmed cases, *C*, and the number of daily recorded new cases *D* would be

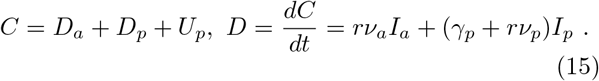

Note that we include *U*_*p*_ because these are people who are not detected through directed tests but eventually get detected (after 1*/γ*_*p*_ days) when they get very sick and go to hospitals. On the other hand the class *D*_*p*_ get detected because of directed testing, even before they get very sick.

### B. Linear analysis of the dynamical equations

Since at early times *S* ≈ *N* and all the other populations *E, I*_*a*_, *I*_*p*_, *D*_*a*_, *D*_*p*_, *U*_*a*_, *U*_*p*_ ≪*N*, one can perform a linearization of the above equations. This tells us about the early time growth of the pandemic, in particular the exponential growth rate. Let us define new variables to characterize the linear regime: *x*_1_ = *S*−*N, x*_2_ = *E, x*_3_ = *I*_*a*_, *x*_4_ = *I*_*p*_, *x*_5_ = *U*_*a*_, *x*_6_ = *D*_*a*_, *x*_7_ = *U*_*p*_, *x*_8_ = *D*_*p*_. At early times when *x*_*i*_ << *N*, the dynamics is captured by linear equations

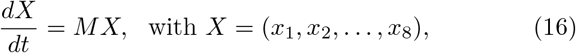

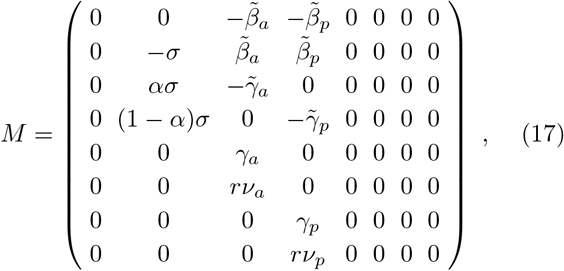

Where 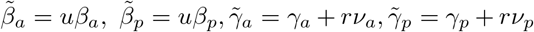. For the present we ignore the time dependence of the SD factor *u* and the TQ factor *r*. The matrix has 5 zero eigenvalues while the 3 non-vanishing ones are given by the roots of the following cubic equation for *λ*:

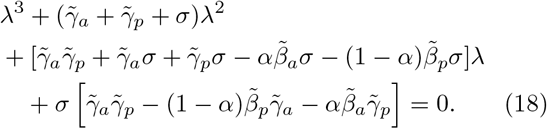

This can be re-written in the form

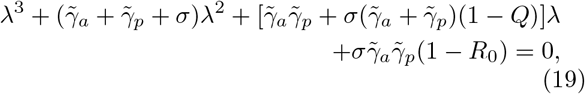

Where 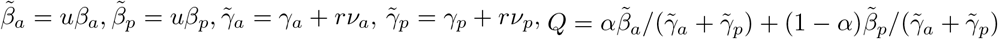 and

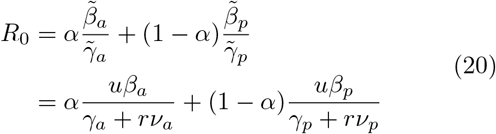

is the expected form for the reproductive number for the disease. Noting the fact that *Q* < *R*_0_, it follows that the condition for at least one positive eigenvalue is

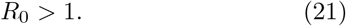

We denote the largest eigenvalue by *µ*. At early times the number of cases detected would grow as ∼ *e*^*µt*^. For *R*_0_ ≈ 1, we expect that the largest eigenvalue is close to zero and from (19) we can read off the value as

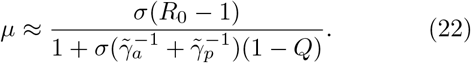

#### Initial conditions

We discuss here the fact that all initial conditions (which satisfy the condition *S*(0) ≈ *N*) quickly move along the direction of the dominant eigen-vector and how this provides us a way to choose the correct initial conditions from the knowledge of one variable (e.g confirmed cases) at an early time. We denote the right and left eigenvectors corresponding to the eigen-value *µ* by *ϕ*_*m*_(*i*) and *χ*_*m*_(*i*) respectively. The time evolution of the vector *X* = (*x*_1_, *x*_2_, *x*_3_, *x*_4_, *x*_5_, *x*_6_, *x*_7_, *x*_8_) is given by

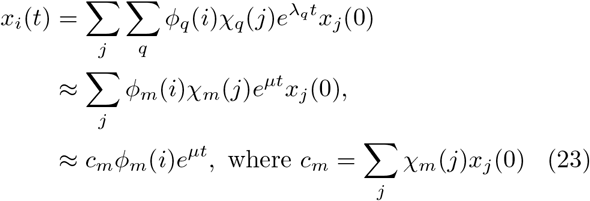

and the second last line is true at sufficiently large times when only one eigenvalue *µ* dominates. This proves that the direction of the vector *X* is independent of initial conditions. In particular, using the explicit form of the dominant eigenvector we find the following relation in the growing phase of the pandemic:

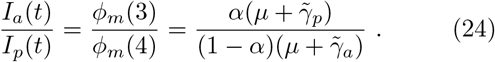

Let us consider the initial condition *X* = (−*E*, 0, 0, *E*, 0, 0, 0, 0) so that (noting that *χ*_*m*_(1) = 0)

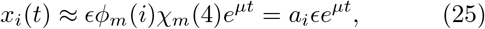

where *a*_*i*_ = *ϕ*_*m*_(*i*)*χ*_*m*_(4). At a sufficiently large time *t*_*l*_ (but still in the very early phase of the pandemic) we equate the observed confirmed number *C*_0_ on some day to *x*_6_(*t*_*l*_) + *x*_7_(*t*_*l*_) + *x*_8_(*t*_*l*_) which therefore gives us the relation

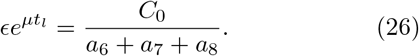

This then tells us that we should start with the following initial conditions, counting now time from *t* = 0 (*i.e*. starting from the day of the observation *C*_0_):

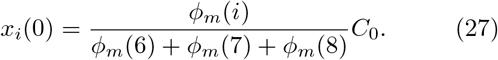

*The crucial point is that the leading eigenvector fixes the direction of the growth and then knowledge of linear combination fixes all the other coordinates*. This also means that trajectories for different initial conditions are identical up to a time translation (See Fig. 4 and related discussion).

### C. Final affected population

Let us define the asymptotic populations (i.e the populations at very long times) in the different compartments as 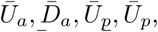, and let 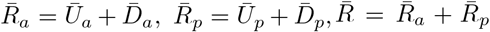. The total population that would eventually be affected by the disease (and either recovered or died) is given by 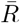 and would have developed immunity. A fraction *Ū a* (see below) would be undetected and uncounted.

It is possible to compute the final affected population 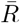 from the dynamical equations in 7 - 14. For the moment let us assume that *u* and *r* do not have any time dependence. We also assume that *U*_*a*_(0) = 0, *U*_*p*_(0) = 0, *D*_*a*_(0) = 0, *D*_*p*_(0) = 0 and *S*(0) ≈ *N*. Then solving (7), we get

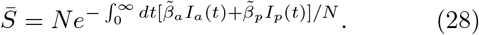

Where 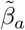 and 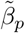 are given after (19). Adding Eqs. (11) and (12) and then multiplying both sides by 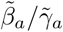 we get 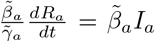 where *R*_*a*_= *U*_*a*_ + *D*_*a*_. Similarly, we also get 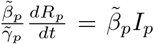 where *R*_*p*_= *U*_*p*_ + *D*_*p*_ Plugging these two equations into (28) then gives

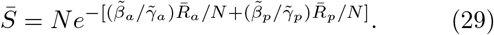

Next we note that (*d/dt*)(*I*_*a*_+*R*_*a*_) = *ασE* and (*d/dt*)(*I*_*p*_+ *R*_*p*_) = (1 − *α*)*σE*. Hence, for the initial condition *I*_*a*_ = *I*_*p*_ = *R*_*a*_ = *R*_*p*_ = 0, we find that the ratio [*I*_*a*_(*t*) + *R*_*a*_(*t*)]*/*[*I*_*p*_(*t*) + *R*_*p*_(*t*)] = *α/*(1 − *α*) at all times. Since at large times *I*_*a,p*_ → 0, this means that the asymptotic values of *R*_*a*_ and *R*_*p*_ are given by

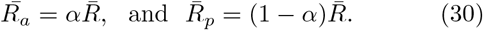

Using this in (29), noting that 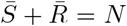 and defining 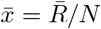, we then get the following simple equation that determines the asymptotic total affected population:

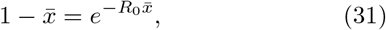

where 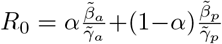 is the reproductive number as stated earlier. We note that (31) has a non-zero solution only when *R*_0_ > 1. For the simple SIR model this result is well known [16], here we show that this is valid for an extended model as well generally. This computation of the asymptotic population can be straightforwardly extended to a more general model where one can have arbitrary number of compartments for the infected and recovered populations.

The asymptotic population of the individual populations are then given by

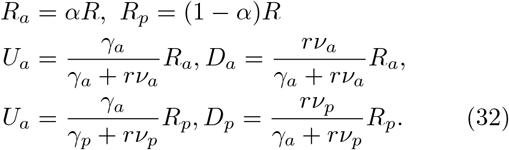

### D. Peak infections and the time at the peak

To compute the maximum of the infected population *I*^(*m*)^ and the time *t*^(*m*)^ at which this peak appears, let us first look at the basic SEIR model consisting of four compartments with populations *S, E, I* and *R*, and the dynamics of these variables described by

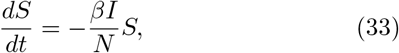

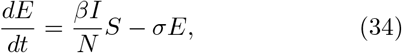

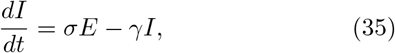

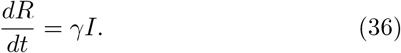

where now *β* is the infectivity, *E* → *I* transitions happens at a rate σ and recovery *I* → *R* happens at a rate *γ*. In this case the reproductive number is simply given by *R*_0_ = *β/γ*. As shown in the previous section it is easy to see that *S*(*t*) and *R*(*t*) are related by 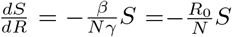 which gives *S*(*t*) = *S*(0)*e*^−*R*^0 *R*(*t*)*/N*. The peak of the infected population is given by setting *dI/dt* = 0 at *t* = *t*^(*m*)^. By solving the equations (33)-(36) for several set of parameters we observe that *E* also achieves its peak around the same time. Hence setting also *dE/dt* = 0 at *t* = *t*^(*m*)^, we find *S*(*t*^(*m*)^) = *S*^(*m*)^*/N* = 1*/R*_0_. On the other hand from *S*(*t*) = *S*(0)*e*^−*R*^0 *R*(*t*)*/N* we get 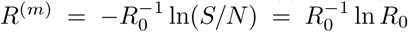. Then using the overall constraint *N* = *S* + *E* + *I* + *R* we finally obtain that the peak value of the infection number is given by

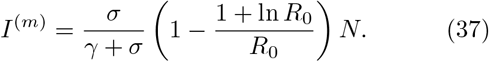

An estimate of the time to reach this peak value can be obtained [17] by noting that we can use the linearized dynamics (see previous section) till the time *I*(*t*) reaches its peak *I*^(*m*)^. Hence we write 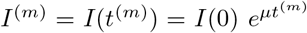 which finally gives

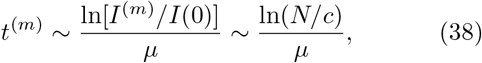

where *c* is a constant that depends on initial infection numbers and parameter values. A verification of this result, obtained by solving the basic SEIR equations numerically, is provided in Fig. 6.

**FIG. 6.**
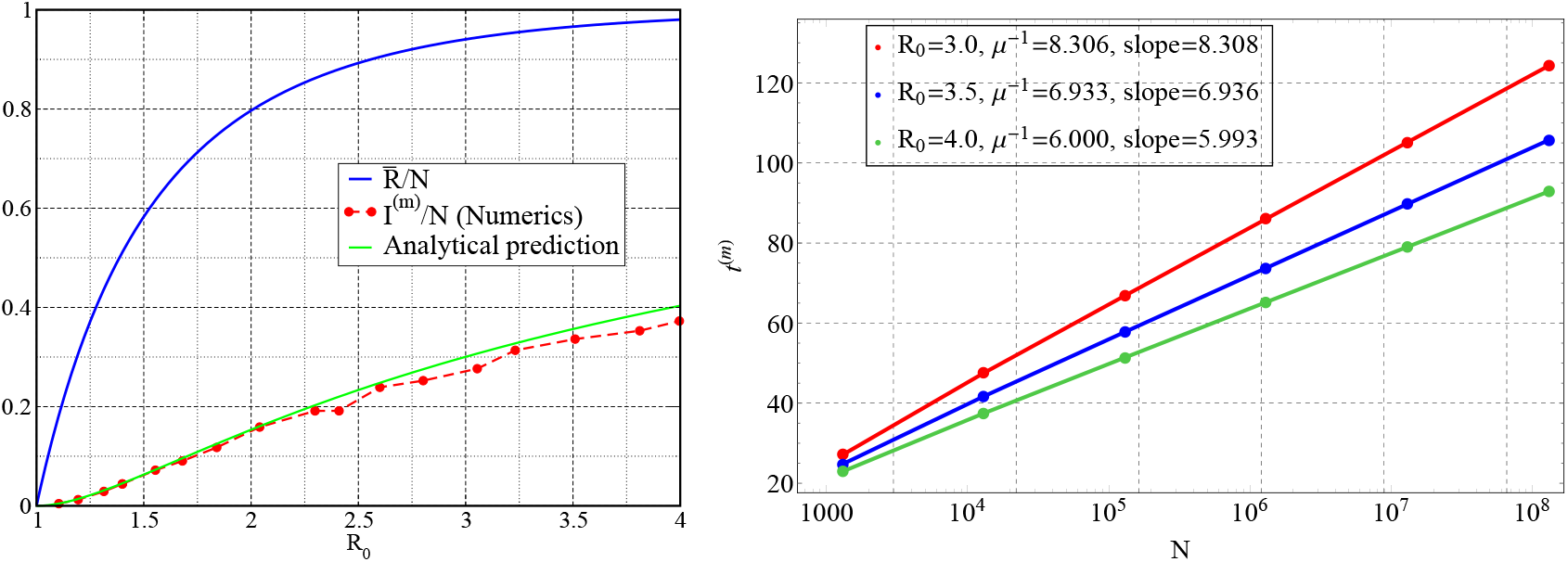
(left) Plot of the asymptotic total affected population fraction, 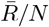, as a function of the reproductive number *R*_0_. We also plot the quantity (*I*^(*m*)^*/N*)(σ+ *γ*_*e*_)*/σ*, obtained numerically from many different parameter sets, and compare it with the theoretical predicted curve 1− (1 + ln *R*_0_)*/R*_0_ (green line). (right) Verification of the ln(*N*) dependence of *t*^(*m*)^ in (38) for different choices of *R*_0_. The slopes of the straight lines compares well with *µ*^−1^ as stated in (38).

Interestingly, we find that the expression for *I*^(*m*)^ also describes quite accurately the peak value for the extended SEIR dynamics with *γ* now replaced by

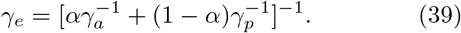

In Fig. 6 we show the dependence of 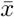 on *R*_0_ (as obtained from a numerical solution of (31)) and provide a numerical verification of the result in Eqs. (37,39) for the extended SEIR model. The peak values of the asymptomatic and presymptomatic populations are respectively given by

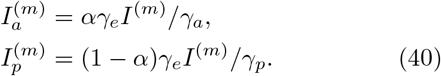

In Fig. 9 we show results of a numerical solution of the dynamical equations in presence of intervention (SD) for one of the parameter sets in Table. III and find excellent agreement with our analytic formula in Eqs. (31,37-39). We see that the predicted peak time is off by about 10%. The numerics also shows that the complete suppression of the disease takes more than 6 months after the peak.

**FIG. 7.**
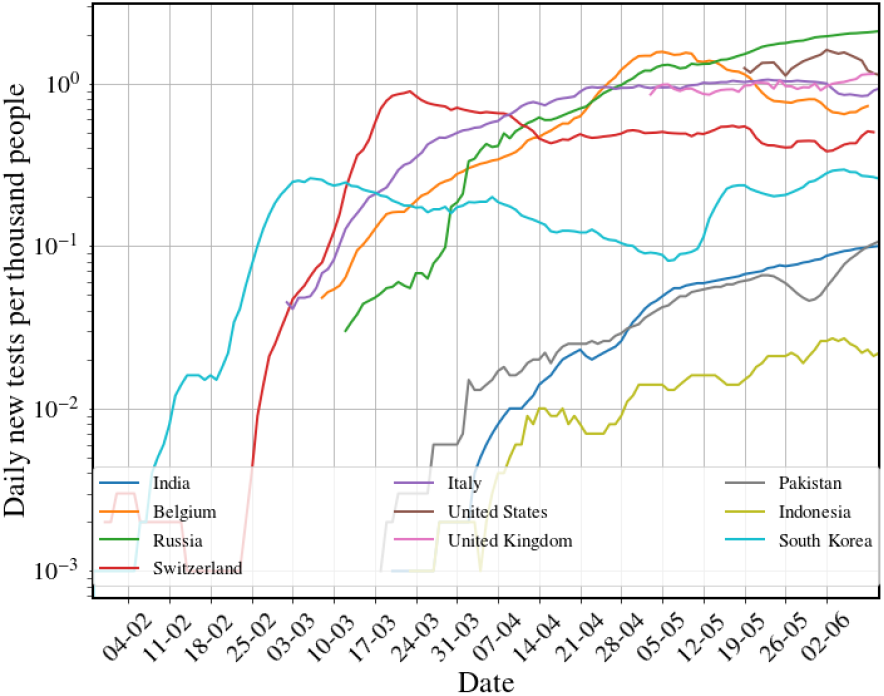
Data of number of tests per day per thousand in several countries on a log-scale. Notice in particular the large testing numbers in the early period in Korea which has been very successful at controlling the disease. Data from [18].

**FIG. 8.**
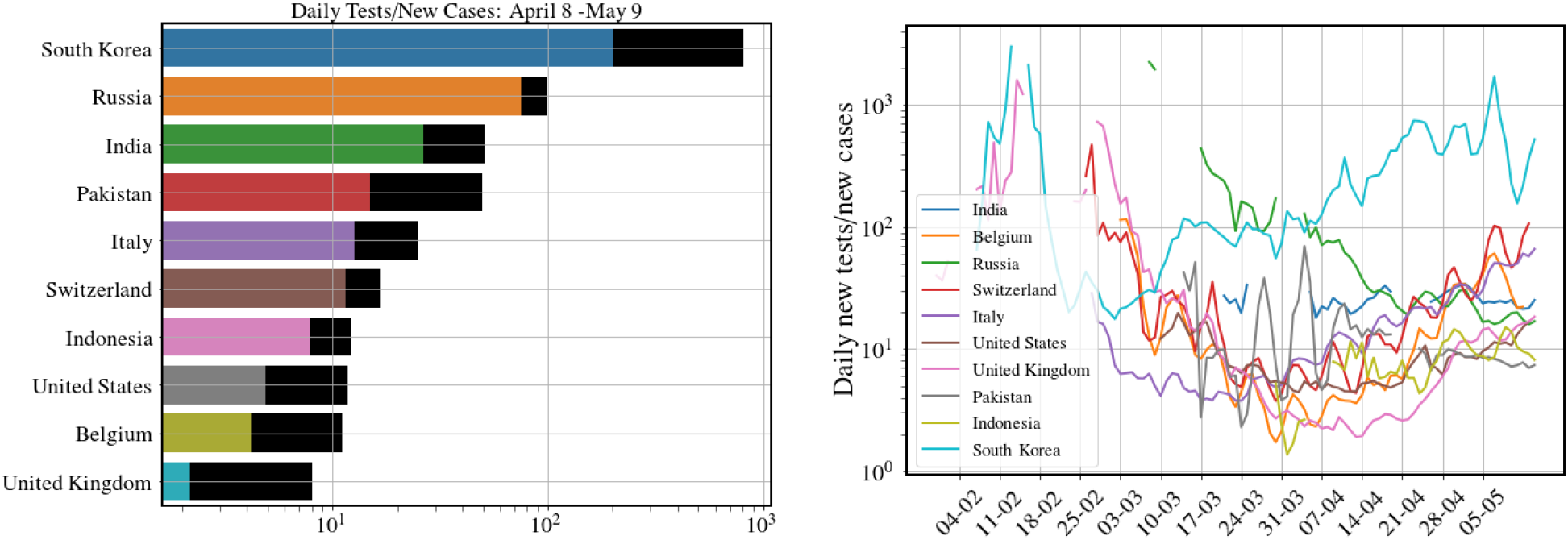
Data from different countries on the number of tests per detected case *T* (*t*)*/D*(*t*) for 10 countries (left) on the dates April 8 (coloured bar), May 9 (black bar) and (right) the change over time of this ratio. Data from [18].

**FIG. 9.**
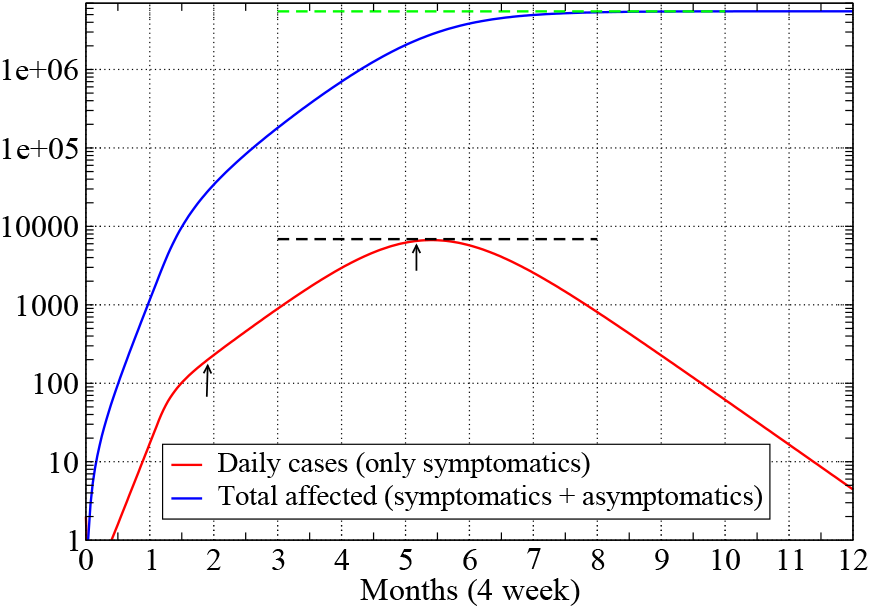
Plot of the new daily cases (*γ*_*p*_*I*_*p*_) and total affected population fraction (*R*), as a function number of months for one of the parameter sets in Table. III for the city of Mumbai. Parameter values were σ= 0.5, 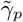, *α* = 0.9, *R*_0_ = 2.16 before intervention and 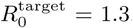. The dashed lines give the analytic predictions for the peak daily cases (black line) and the final affected population (green line), and show the good agreement with the numerics. The arrows indicate the date when initial condition was specified *γ*_*p*_*I*_*p*_(0) = 195 and the peak infection date, which occurs about 15 days after the date predicted from (38).

### E. Interventions: Social distancing and Testing-Quarantining

We discuss here the choices of the intervention functions *u* and *r* introduced in the dynamical equations 7-14. Note that *u* is a dimensionless number quantifying the level of social contacts, while *r* is a rate which, as we will see, is closely related to the testing rate.

#### Social distancing (SD)

We multiply the constant factors *β*_*a,p*_ by the time dependent function, *u*(*t*), the “lockdown” function that incorporates the effect of a social distancing, i.e reducing contacts between people. A reasonable form is one where *u*(*t*) has the constant value (= 1) before the beginning of any interventions, and then from time *t*_*on*_ it changes to a value 0 < *u*_*l*_ < 1, over a characteristic time scale ∼ *t*_*w*_. Thus we take a form

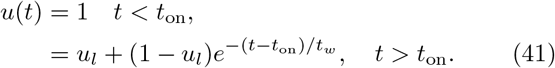

The number *u*_*l*_ indicates the lowering of social contacts.

#### Testing-quarantining (TQ)

We expect that testing and quarantining will take out individuals from the infectious population and this is captured by the terms *rν*_*a*_*I*_*a*_ and *rν*_*p*_*I*_*p*_ in the dynamical equations. A reasonable choice for the TQ function is perhaps to take

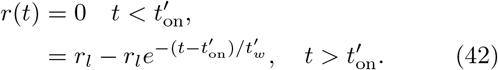

where we one needs a final rate *r*_*l*_ > 0. In general the time at which the TQ begins to be implemented 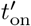 and the time required for it to be effective 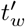 could be different from those used for SD.

A useful quantity to characterize the system with interventions is the time-dependent effective reproductive number given by

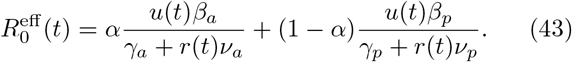

At long times this goes to the targeted reproduction number

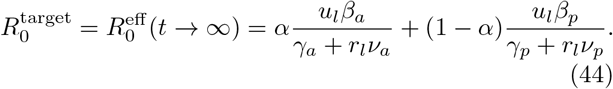

The time scale for the intervention target to be achieved is given by *t*_*w*_ and *t*^*l*^

#### Relation of the TQ function *r*(*t*) to the number of tests done per day

Let us suppose that the number of tests per person per day is given by *T*_*r*_. We show in Fig. (7) the data for the number of tests per 1000 people per day across a set of countries and see that this is around 0.05 for India which means that *T*_*r*_ = 0.00005. If tests are done completely randomly, then the number of detected people (assuming that the tests are perfect) would be *T*_*r*_ ×*I* and so it is clear that we can identify *r*(*t*) = *T*_*r*_(*t*). It is then clear that this would have no effect on the pandemic control. To have any effect we would need *r γ*_*p*_≈0.1 which means around 100 tests per 1000 people per day which is clearly not practical.

However, a better strategy is to do focused tests on the contacts of all those who have been detected on a given day. We now give an estimate of the rate *r* if we followed this strategy. For simplicity of presentation of our argument we here assume *ν*_*a*_ = *ν*_*p*_ = 1 and *γ*_*a*_ = *γ*_*p*_. From our extended SEIR model the number of detected cases per day is given by Δ*D*(*t*) = *rν*_*a*_*I*_*a*_ +(*rν*_*p*_ + *γ*_*p*_)*I*_*p*_ = *rI* + *γ*_*p*_*I*_*p*_. In the growing phase we have, from Eq. (24), that *I*_*a*_ = *αI* and *I*_*p*_ = (1 −*α*)*I*. Hence we get 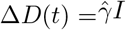 with 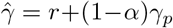. The total number of contacts of the 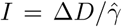 individuals would be 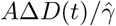, where *A* is the mean number of contacts of a single infected person. If we perform *T* tests per day *on this pool*, then the rate of detections will be given by

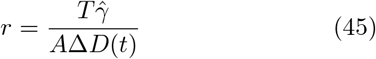

Denoting *c* = *T/*(*A*Δ*D*) and noting that 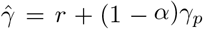, we self-consistently solve the above equation to find

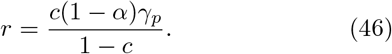

Now it is clear that unless *r* and *γ*_*a*_ = *γ*_*p*_ are of the same order, TQ will not have much effect on the dynamics and the change in *R*_0_ will be small. Setting *r γ*_*p*_ then gives us the condition

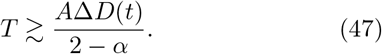

Note that in our model we identify *r*(*t*) as our control rate function that changes from the value 0 to a value *r*_*l*_ ≈ *γ*_*p*_ over the time scales of a week or so. This means that we would need to change the testing rate in a controlled way such that the condition *T* (*t*) ∼ *A*Δ*D*(*t*) is maintained. The implications of this is discussed after (6). In Fig. 7 we show data for daily new tests for a set of countries. A noteworthy case is the data for South Korea where we see the large testing rate at early days of the pandemic. Perhaps this explains the quick control of the pandemic in that country. The table in Fig. 8 shows data for the ratio *T* (*t*)*/*Δ*D*(*t*) for a set of countries and also how this ratio has evolved over time. As explained earlier (see Sec. I C), the number *A* is expected to depend on the population density and also how well SD is being implemented and hence, for a country like India *T* (*t*)*/*Δ*D*(*t*) ≈ 25 may not be sufficient.

## ACKNOWLEDGMENTS

We thank Jitendra Kethepalli and Kanaya Malakar for very helpful discussions and Ranjini Bandyopadhyay, Siddhartha Chatterjee, Joel Lebowitz, Srujana Merugu, Alpan Raval, Sankar Das Sarma and Sriram Shastry for a careful reading of the draft and making useful suggestions. We acknowledge support of the Department of Atomic Energy, Government of India, under project no.12-R&D-TFR-5.10-1100.

## AUTHOR CONTRIBUTIONS

All authors performed research; S. Goyal analyzed data; A. Das, A. Dhar, A. Kundu wrote the paper.

## References

[1] Kucharski, A. J. et al. Early dynamics of transmission and control of COVID-19: a mathematical modelling study. Lancet Infect Dis 20, 533–38 (2020) https://doi.org/10.1016/S1473-3099(20)30144-4.

[2] Prem K., Liu Y,, Russell T. W. et al. The effect of control strategies to reduce social mixing on outcomes of the COVID-19 epidemic in Wuhan, China: a modelling study. Lancet Public Health. 2020; (published online March 25.) https://doi.org/10.1016/S2468-2667(20)30073-6.

[3] Ferguson, N. et al. Report 9:Impact of non-pharmaceutical interventions (NPIs) to reduce COVID19 mortality and healthcare demand. https://doi.org/10.25561/77482 (2020).

[4] Tang, B. et al. An updated estimation of the risk of transmission of the novel coronavirus (2019-nCov)., Infectious Disease Modelling 5, 248 (2020).

[5] Giordano, G. et al. Modelling the COVID-19 epidemic and implementation of population-wide interventions in Italy. Nature Medicine https://doi.org/10.1038/s41591-020-0883-7 (2020).

[6] Gatto, M. et al. Spread and dynamics of the COVID-19 epidemic in Italy: Effects of emergency containment measures. Proceedings of the National Academy of Sciences, 202004978. DOI:10.1073/pnas.2004978117.

[7] Ray, D. et al. Predictions, role of interventions and effects of a historic national lockdown in India’s response to the COVID-19 pandemic: data science call to arms. medRxiv https://doi.org/10.1101/2020.04.15.20067256 (2020).

[8] Sarkar, K. and Khajanchi, S. Modeling and forecasting of the COVID-19 pandemic in India. 2005.07071 (2020).

[9] Shekatkar, S. et al. INDSCI-SIM: A state-level epidemiological model for India. https://indscicov.in/indscisim.

[10] Agrawal, S. et al., COVID-19 Epidemic: Un-locking the lockdown in India (working paper), IISc-TIFR Technical Report, https://covid19.iisc.ac.in/wp-content/uploads/2020/04/Report-1-20200419-UnlockingTheLockdownInIndia.pdf.

[11] Venkateswaran, J. Damani, O. Effectiveness of Testing, Tracing, Social Distancing and Hygiene in Tackling Covid-19 in India: A System Dynamics Model, arXiv :2004.08859 (2020).

[12] Meidan, D. et al., Alternating quarantine for sustainable mitigation of COVID-19, arxiv.org:2004.01453 (2020).

[13] George N. Wong et al, 2006.02036 (2020)

[14] Liu, Y., Gayle, A. A., Wilder-Smith, A., Rocklöv J. The reproductive number of COVID-19 is higher compared to SARS coronavirus, Journal of Travel Medicine 27, Issue 2, March 2020, https://doi.org/10.1093/jtm/taaa021.

[15] Leung, K. Y., Trapman, P., Britton, T., Who is the infector? Epidemic models with symptomatic and asymptomatic cases, Mathematical Biosciences, 301, 190 (2018).

[16] Brauer, F., Driessche, P. V., and Wu, J. Lecture notes in Mathematical Epidemiology, Springer (2008).

[17] Li J. and Lou Y., Characteristics of an epidemic out-break with a large initial infection size, Journal of biological dynamics 10, 366 (2016). https://doi.org/10.1080/17513758.2016.1205223

[18] Coronavirus (COVID-19) Testing, https://ourworldindata.org/coronavirus-testing.

[19] Johns Hopkins Center for Systems Science and Engineering Coronavirus COVID-19 Global Cases. https://systems.jhu.edu/research/public-health/ncov/

[20] Data for India and Delhi are from the website https://www.kaggle.com/sudalairajkumar/covid19-in-india/data#covid_19_india.csv/. The numbers for Mumbai are estimated from newspaper reports.

